# COVID-19 and Indian population: a comparative genetic analysis

**DOI:** 10.1101/2021.12.15.21267816

**Authors:** Saurabh Patil, Sandhya Kiran Pemmasani, Neelima Chitturi, Ishita Bhatnagar, Anuradha Acharya, Lingareddy Venkata Subash

## Abstract

**Background:** Major risk factors of COVID-19 include older age, male gender, and comorbidities. In addition, host genetic makeup is also known to play a major role in COVID-19 susceptibility and severity. To assess the genetic predisposition of the Indian population to COVID-19, a comparative analysis of the frequencies of polymorphisms directly or potentially associated with COVID-19 susceptibility, severity, immune response, and fatal outcomes was done between the Indian population and other major populations (European, African, East Asian, South Asian, and American).

**Materials and methods:** Polymorphisms directly or potentially associated with COVID-19 susceptibility, severity, immune response, and mortality were mined from genetic association studies, comparative genetic studies, expression quantitative trait loci studies among others. Genotype data of these polymorphisms were either sourced from the GenomegaDB™ database of Mapmygenome India Ltd. (sample size = 3054; Indian origin) or were imputed. Polymorphisms with minor allele frequency >= 0.05 and that are in Hardy-Weinberg equilibrium in the Indian population were considered for allele frequency comparison between the Indian population and 1000 Genome population groups.

**Results:** Allele frequencies of 421 polymorphisms were found to be significantly different in the Indian population compared to European, African, East Asian, South Asian, and American populations. 128 polymorphisms were shortlisted based on linkage disequilibrium and were analyzed in detail. Apart from well-studied genes, like ACE2, TMPRSS2, ADAM17, and FURIN, variants from AHSG, IFITM3, PTPN2, CD209, CCL5, HEATR9, SELENBP9, AGO1, HLA-G, MX1, ICAM3, MUC5B, CRP, C1GALT1, and other genes were also found to be significantly different in Indian population. These variants might be implicated in COVID-19 susceptibility and progression.

**Conclusion:** Our comparative study unraveled multiple genetic variants whose allele frequencies were significantly different in the Indian population and might have a potential role in COVID-19 susceptibility, its severity, and fatal outcomes. This study can be very useful for selecting candidate genes/variants for future COVID-19 related genetic association studies.

## 1 INTRODUCTION

Coronavirus disease - 2019 (COVID-19), caused by severe acute respiratory syndrome coronavirus 2 (SARS-CoV2), has strained the healthcare system and economy of a majority of nations and has crippled the human population [1, 2, 3]. COVID-19 is characterized by a range of clinical features which are categorized as most common, less common, and rare (severe). The most common symptoms are fever, dry cough, and fatigue, while less common symptoms are pneumonia without noticeable hypoxemia, sputum production, sore throat, headache, chest pain, and diarrhea [4, 5]. Acute respiratory distress syndrome, sepsis, acute cardiac injury, heart failure, acute kidney injury, hypoxic encephalopathy are more common in severe cases [4, 5, 6]. Severe patients show elevated levels of pro-inflammatory cytokines, indicating the presence of cytokine storm [4]. Self-reported loss of smell and taste was also observed in some cases [7].

Major risk factors for COVID-19 severity are older age, male gender, and comorbidities. A higher case fatality rate (CFR) has been observed in older adults as compared to the younger population [8, 9, 10, 11, 12]. Relatively fewer female deaths are consistently observed as compared to males [10, 11, 12]. COVID-19 patients with comorbidities like hypertension, obesity, diabetes, and others are more likely to develop severe complications than ones without any underlying diseases [13, 14, 15, 16, 17, 18]. ABO blood groups also seem to be contributing to COVID-19 related risk. While blood group A is associated with an increased risk of severe COVID-19 outcomes, blood group O seems to be protective [19, 20, 21].

In addition to the above risk factors, host genetics can modulate susceptibility and severity of the disease by regulating viral entry and host immune response [22, 23]. For example, rs12329760 (p.Val197Met) affects the stability of TMPRSS2 protein, which is implicated in SARS-CoV2 viral entry into host cells. This polymorphism was found to be less frequent in severe COVID-19 cases as compared to others, suggesting its protective role against severe outcomes of COVID-19 [24]. The C allele of rs12252 in the IFITM3 gene that is known for inhibiting influenza virus entry into host cells was found to be a genetic risk factor for COVID-19 hospitalization and severe outcomes [25, 26]. 3p21.31 gene cluster confers genetic susceptibility to COVID-19 with respiratory failure in Italian and Spanish populations [27]. Genetic variants present in immune-related genes like HLA [24, 28, 29, 30], IL-6 [24], TNF-alpha [31], C5 [32, 33] are associated with COVID-19 severity.

While the majority of these studies were done on European and East Asian populations, limited studies were done on the Indian population [34, 35, 36]. Considering the significant role of host genetics in COVID-19 susceptibility and severity in the Indian population, we analyzed the frequencies of the polymorphisms that are directly or indirectly implicated in COVID-19. We have also made a comparison between Indian and other populations to understand allele frequency distribution globally.

## 2 MATERIALS AND METHODS

### 2.1 Literature mining and polymorphism selection

Research articles related to COVID-19 genetic associations were searched in Pubmed using keywords such as ‘COVID’, ‘COVID-19’, ‘Corona virus’, ‘GWAS’, ‘genetic susceptibility’, ‘polymorphism’, ‘severity’, ‘susceptibility’ among other relevant terms. A total of 1418 research articles were found, which were thoroughly screened and 99 relevant articles were selected. A total of 1974 polymorphisms were shortlisted from the selected articles.

### 2.2 Genotype data for Indian population

The GenomegaDB™ database of Mapmygenome India Ltd. [153] was accessed to obtain genotype data for the present study. It is a human polymorphism genotype database developed and owned by Mapmygenome. It contains genotype data of more than 20000 individuals of various ethnicities. Genotype data of 3035 individuals with confirmed Indian origin were considered in this study. For polymorphisms not present in the database, genotypes were imputed with IMPUTE2 using 1000 Genomes Phase3 data as reference [37, 38]. Filtration was performed with an imputation certainty score (Info metric) set at 0.3.

### 2.3 Genotype data for other populations

Allelic and genotypic frequency data of selected polymorphisms for African (AFR), East Asian (EAS), European (EUR), South Asian (SAS), and American (AMR) populations were taken from 1000 Genomes Phase 3 data [38].

### 2.4 Statistical analysis

Polymorphisms having minor allele frequency (MAF) greater than 0.05 in the Indian population (study population) and are in Hardy-Weinberg equilibrium (HWE) (p>0.001) in the Indian population were considered. Pearson’s chi-square test was conducted to compare allele frequencies between the Indian population and other populations (AFR, AMR, EUR, EAS, SAS). False discovery rate (FDR) was applied to correct for multiple comparisons. These statistical tests were done with the R packages HardyWeinberg and stats [39]. LDLink tool [40] was used to calculate linkage disequilibrium (LD) among the polymorphisms.

### 2.5 eQTL data

To analyze the effect of polymorphisms on gene expressions, expression quantitative trait loci (eQTL) data from publicly available GTEx Project portal was used [41]

## 3 RESULTS

Out of 1974 shortlisted polymorphisms, 936 polymorphisms had genotype information, either from GenomegaDB™ or from imputation. Among them, 670 polymorphisms that had MAF>=0.05 in the Indian population were considered for further analysis. Followed by this, 589 polymorphisms that were in HWE (p>0.001) were selected. Their allele frequencies in the Indian population were compared with those in other populations using Pearson’s chi-square test of significance (Supplementary File 1, Supplementary Table 1). 421 polymorphisms had significant differences (FDR <= 0.01) in at least three populations with respect to the Indian population (Supplementary File 1, Supplementary Table 2). To rule out redundancy, LD blocks were obtained, and finally, 128 representative polymorphisms were considered to discuss in detail (Supplementary File 2).

Polymorphisms were categorized based on their effect on susceptibility to COVID-19, immune response to SARS-CoV2 infection, COVID-19 severity, mortality related to COVID-19, and comorbidities. Fifty polymorphisms directly or indirectly associated with COVID-19 susceptibility (Table 1), eighteen polymorphisms implicated in the immune response to SARS-CoV2 infection (Table 2), fifty-eight polymorphisms directly or indirectly associated with COVID-19 severity (Table 3), nine polymorphisms associated with COVID-19 related mortality/fatal outcomes (Table 4), and twelve polymorphisms associated with comorbidities of COVID-19 (Table 5) were analyzed.

**Table 1:**
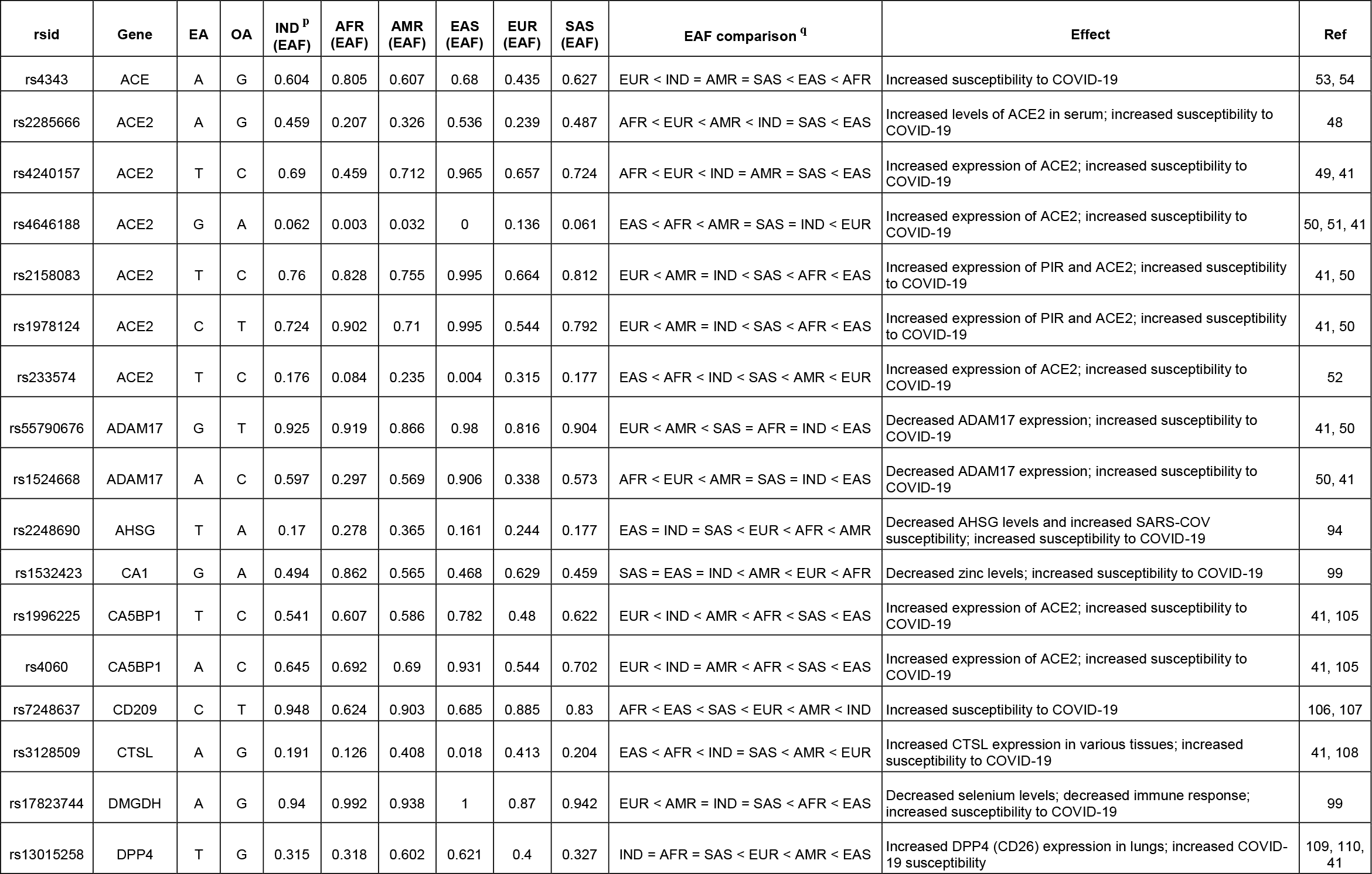
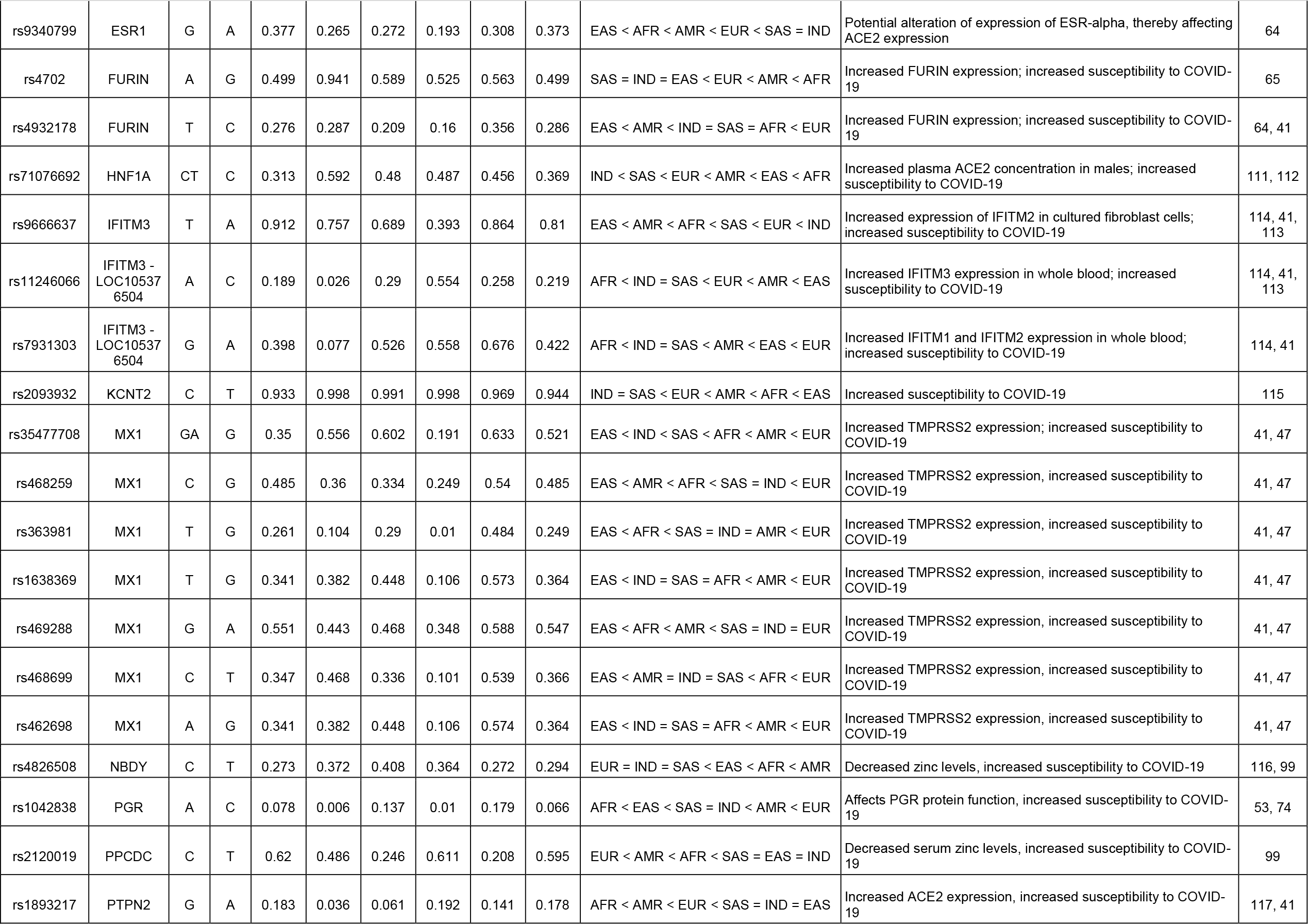
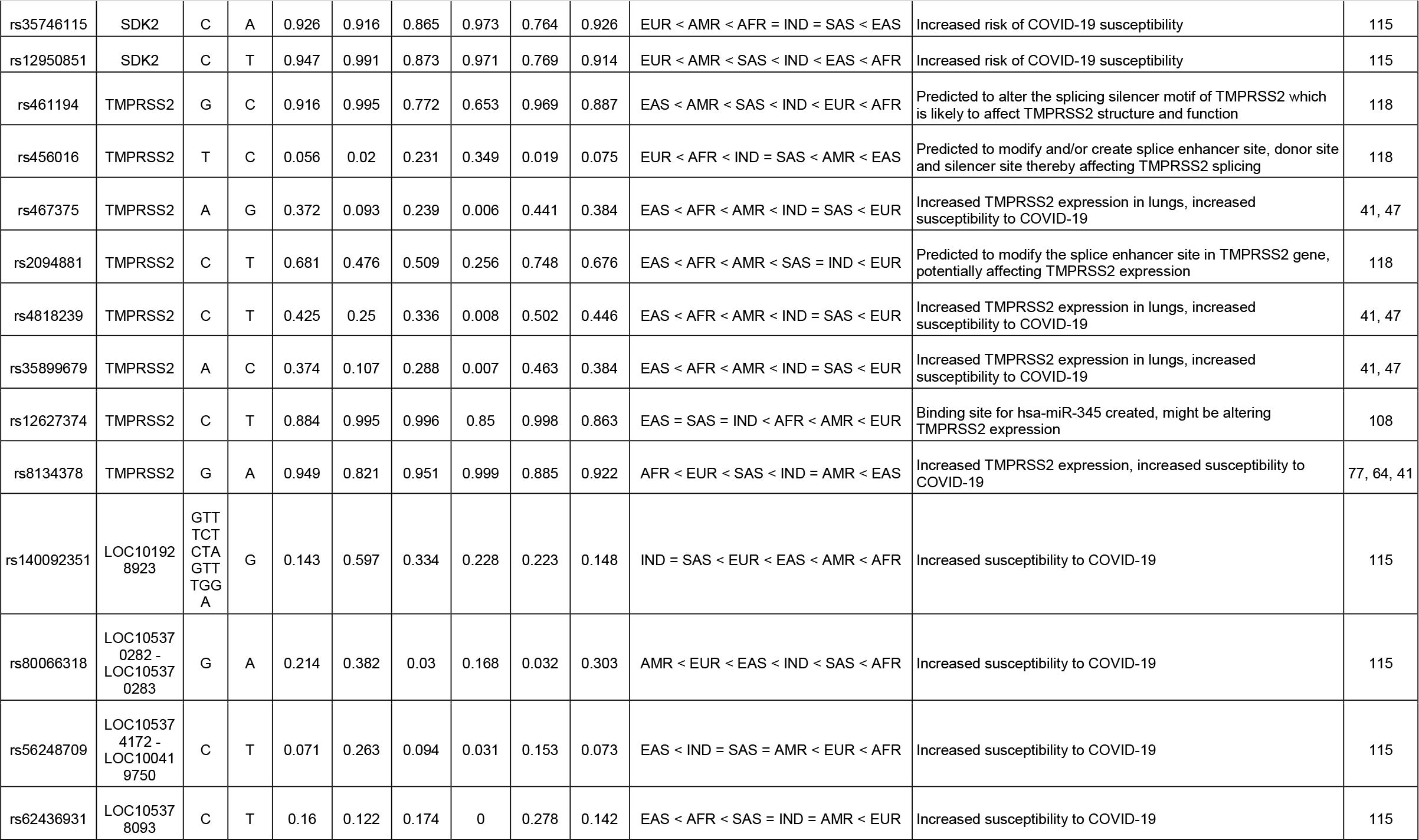
Genetic variants related to COVID-19 susceptibility. ^p^ Effect allele frequency in Indian population. ^q^ EAF (India) compared with EAF in other populations (IND – India, EUR – Europe, AMR – America, SAS – South Asia, AFR – Africa, EAS – East Asia. ‘=’ denotes no significant difference between EAF of Indian and the other population. (EA – effect allele, OA – other allele, Ref – references).

**Table 2:**
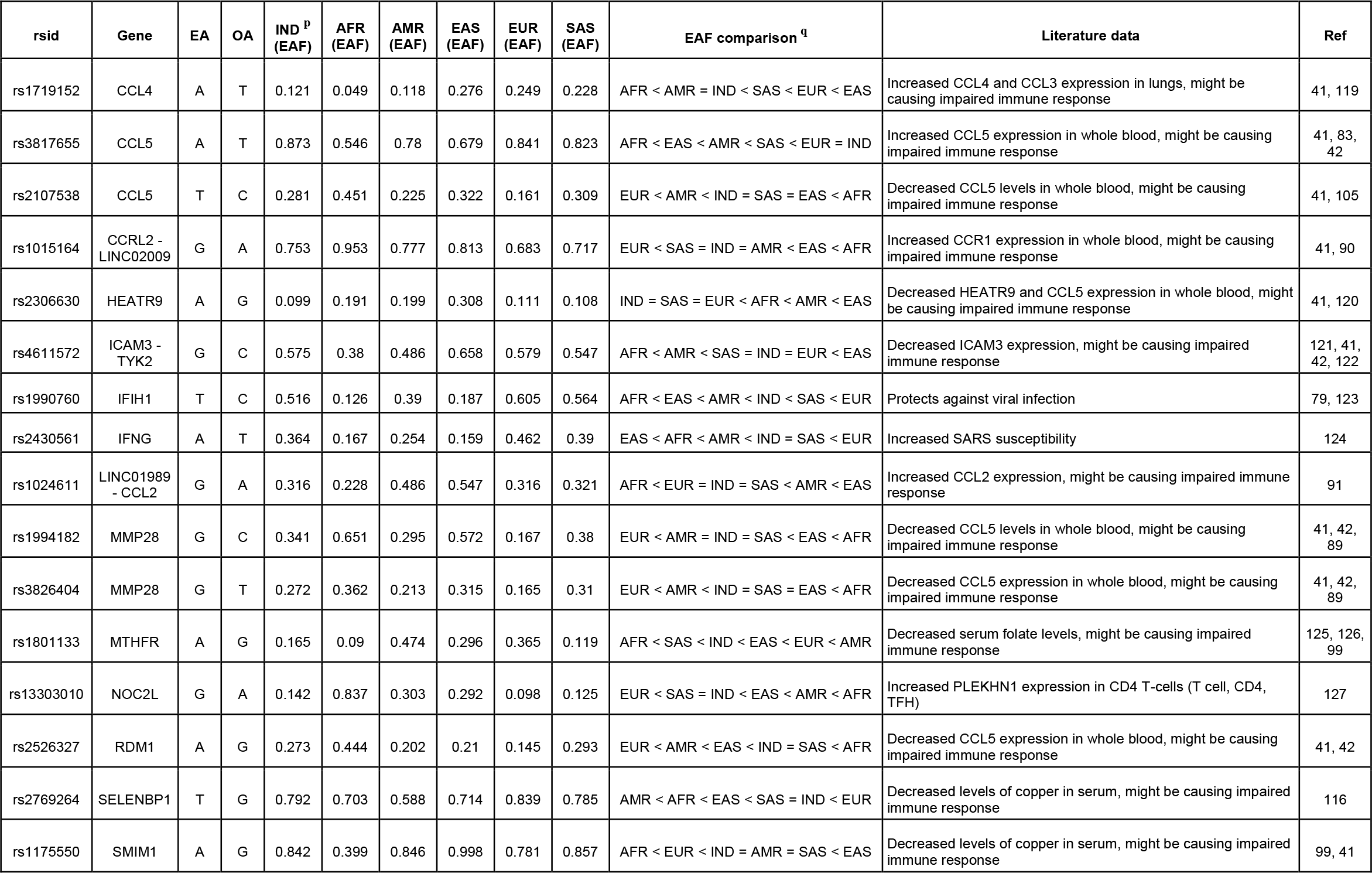
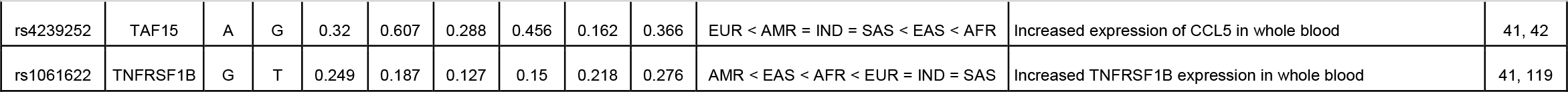
Genetic variants related to host immune response to SARS-CoV2 infection. ^p^ Effect allele frequency in Indian population. ^q^ EAF (India) compared with EAF in other populations (IND – India, EUR – Europe, AMR – America, SAS – South Asia, AFR – Africa, EAS – East Asia. ‘=’ denotes no significant difference between EAF of Indian and the other population. (EA – effect allele, OA – other allele, Ref – references).

**Table 3:**
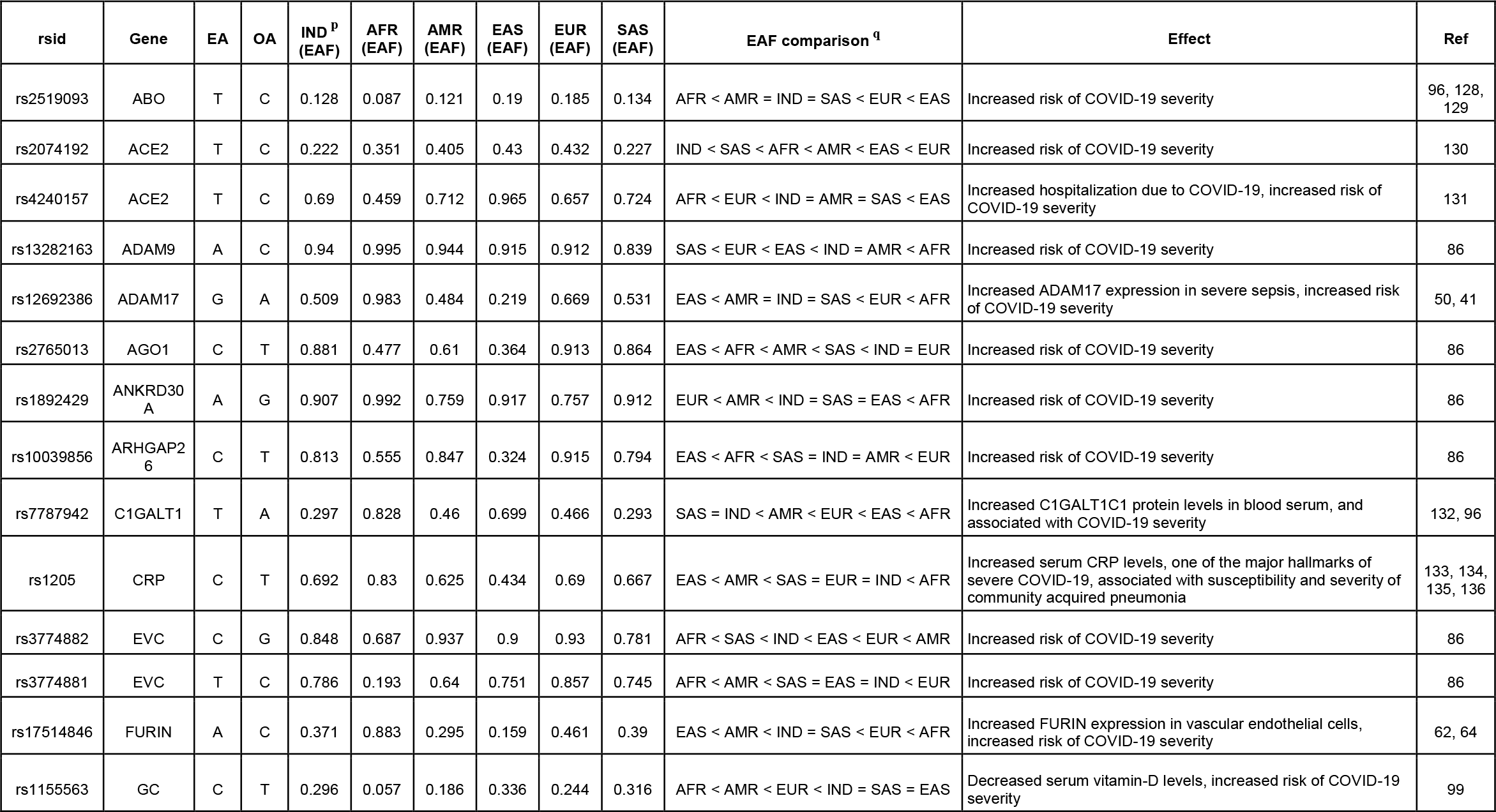
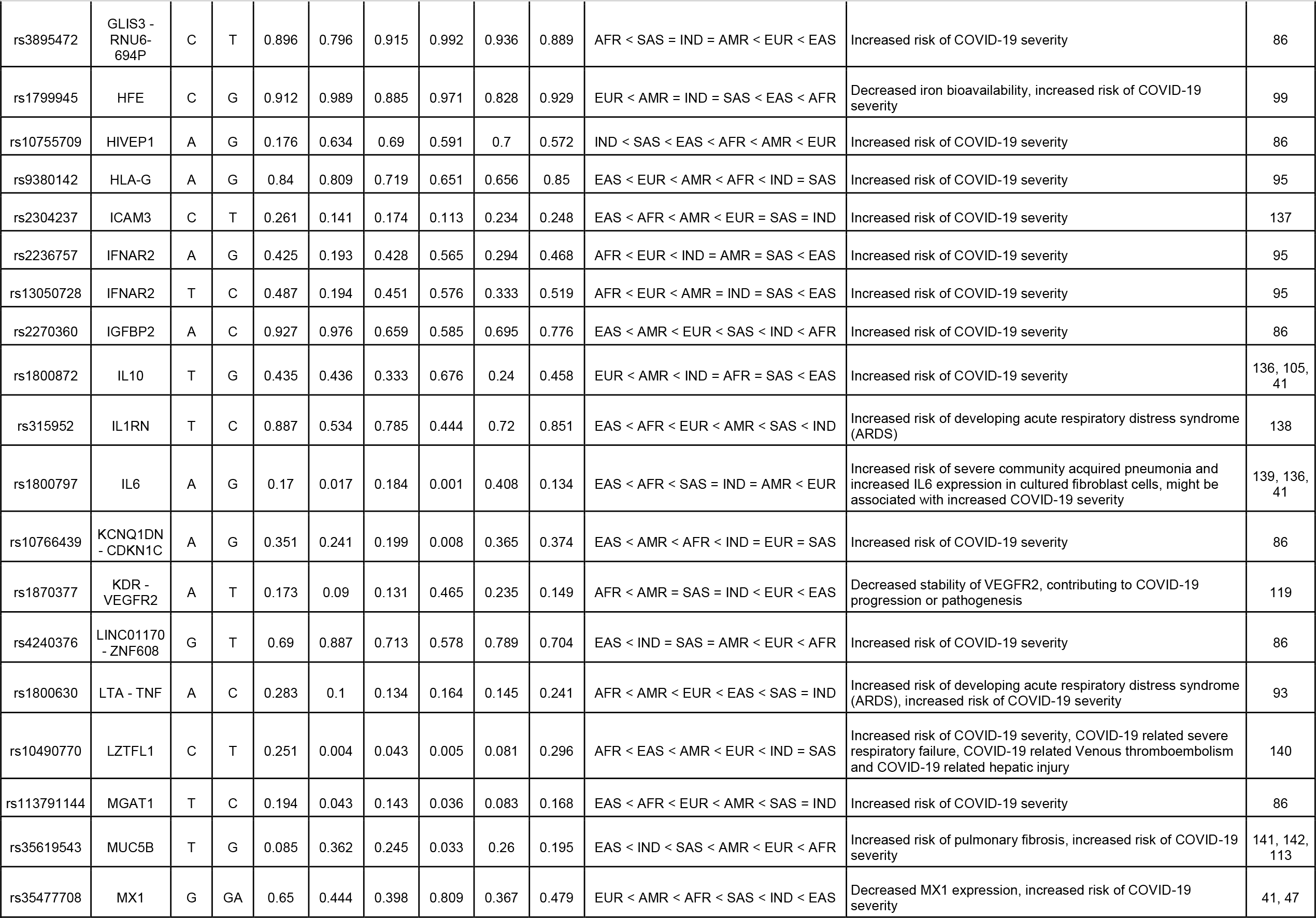
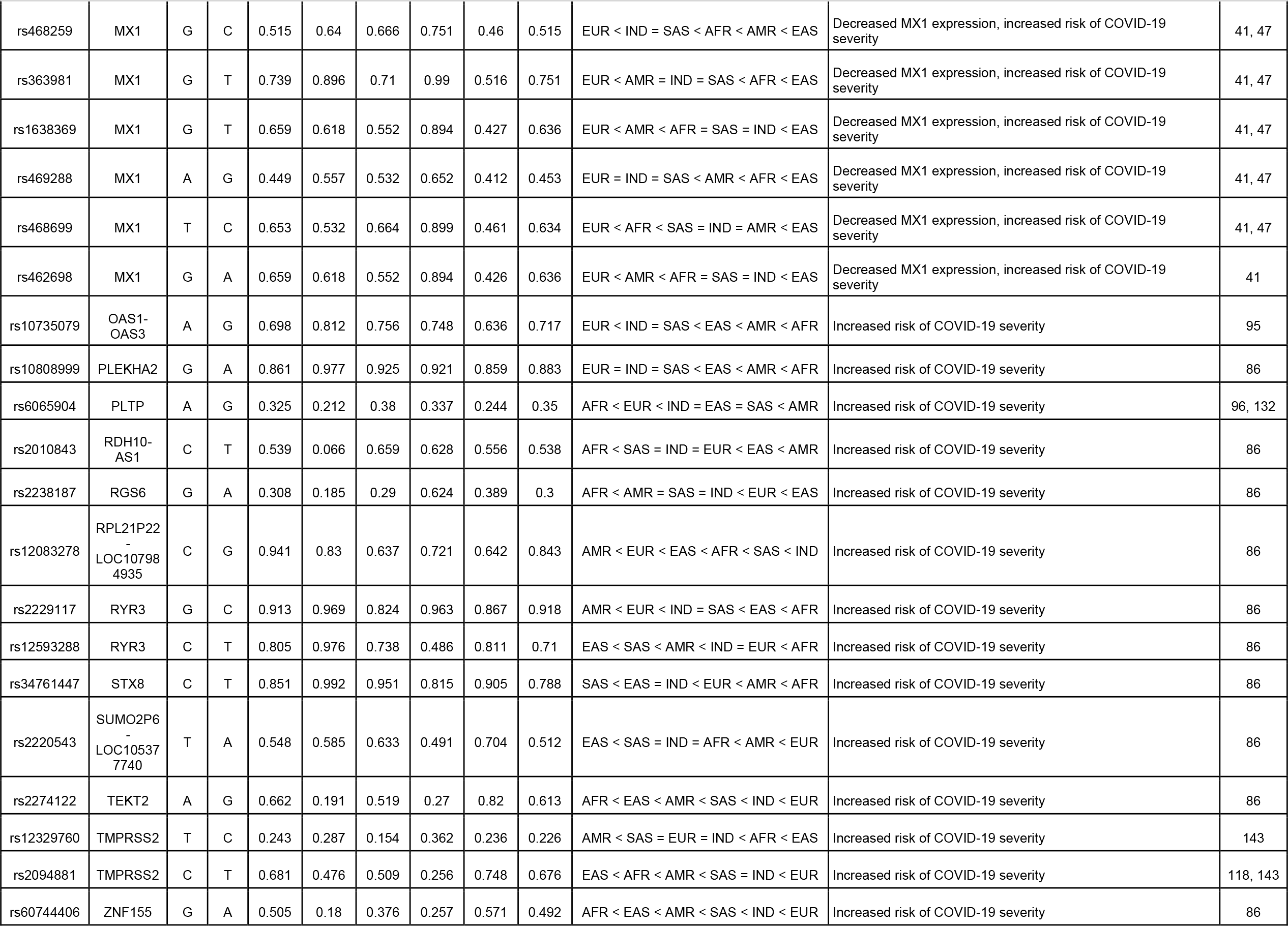
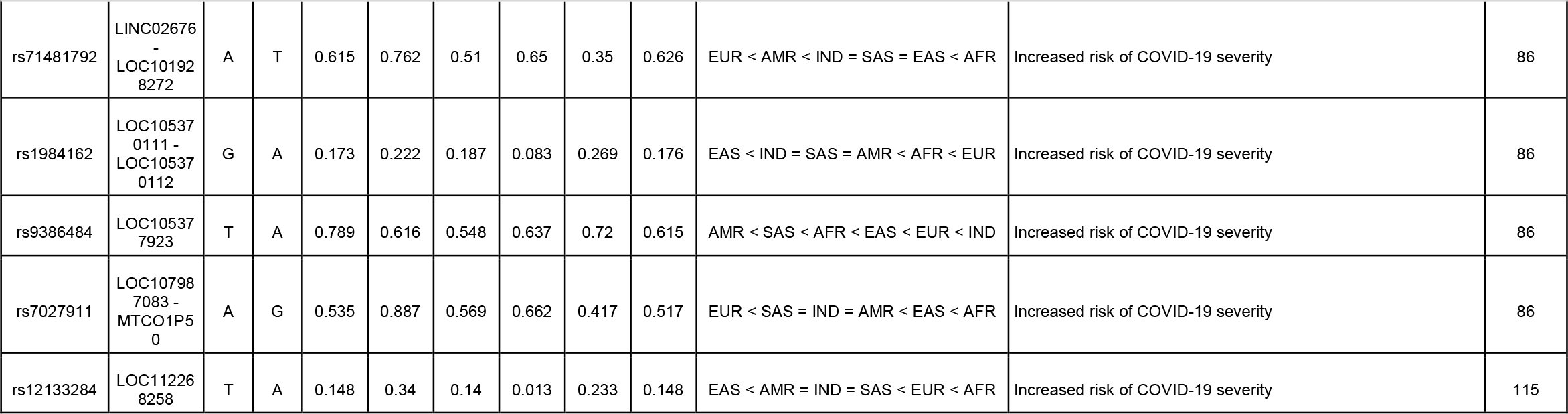
Genetic variants related to COVID-19 severity. ^p^ Effect allele frequency in Indian population. ^q^ EAF (India) compared with EAF in other populations (IND – India, EUR – Europe, AMR – America, SAS – South Asia, AFR – Africa, EAS – East Asia. ‘=’ denotes no significant difference between EAF of Indian and the other population. (EA – effect allele, OA – other allele, Ref – references).

**Table 4:**
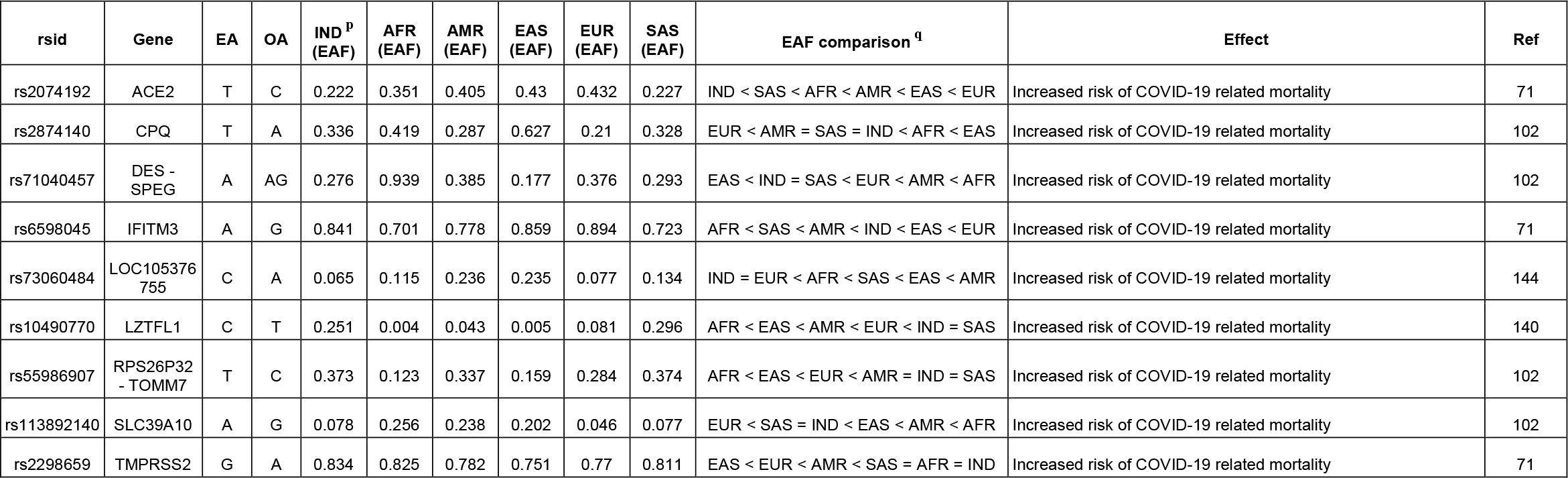
Genetic variants related to COVID-19 related fatal outcomes or mortality. ^p^ Effect allele frequency in Indian population. ^q^ EAF (India) compared with EAF in other populations (IND – India, EUR – Europe, AMR – America, SAS – South Asia, AFR – Africa, EAS – East Asia. ‘=’ denotes no significant difference between EAF of Indian and the other population. (EA – effect allele, OA – other allele, Ref – references).

| rsid | Gene | EA | OA | IND <sup>p</sup> (EAF) | AFR (EAF) | AMR (EAF) | EAS (EAF) | EUR (EAF) | SAS (EAF) | EAF comparison <sup>q</sup> | Effect | Ref |
| --- | --- | --- | --- | --- | --- | --- | --- | --- | --- | --- | --- | --- |
| rs2074192 | ACE2 | T | C | 0.222 | 0.351 | 0.405 | 0.43 | 0.432 | 0.227 | IND < SAS < AFR < AMR < EAS < EUR | Increased risk of COVID-19 related mortality | 71 |
| rs2874140 | CPQ | T | A | 0.336 | 0.419 | 0.287 | 0.627 | 0.21 | 0.328 | EUR < AMR = SAS = IND < AFR < EAS | Increased risk of COVID-19 related mortality | 102 |
| rs71040457 | DES - SPEG | A | AG | 0.276 | 0.939 | 0.385 | 0.177 | 0.376 | 0.293 | EAS < IND = SAS < EUR < AMR < AFR | Increased risk of COVID-19 related mortality | 102 |
| rs6598045 | IFITM3 | A | G | 0.841 | 0.701 | 0.778 | 0.859 | 0.894 | 0.723 | AFR < SAS < AMR < IND < EAS < EUR | Increased risk of COVID-19 related mortality | 71 |
| rs73060484 | LOC105376755 | C | A | 0.065 | 0.115 | 0.236 | 0.235 | 0.077 | 0.134 | IND = EUR < AFR < SAS < EAS < AMR | Increased risk of COVID-19 related mortality | 144 |
| rs10490770 | LZTFL1 | C | T | 0.251 | 0.004 | 0.043 | 0.005 | 0.081 | 0.296 | AFR < EAS < AMR < EUR < IND = SAS | Increased risk of COVID-19 related mortality | 140 |
| rs55986907 | RPS26P32 - TOMM7 | T | C | 0.373 | 0.123 | 0.337 | 0.159 | 0.284 | 0.374 | AFR < EAS < EUR < AMR = IND = SAS | Increased risk of COVID-19 related mortality | 102 |
| rs113892140 | SLC39A10 | A | G | 0.078 | 0.256 | 0.238 | 0.202 | 0.046 | 0.077 | EUR < SAS = IND < EAS < AMR < AFR | Increased risk of COVID-19 related mortality | 102 |
| rs2298659 | TMPRSS2 | G | A | 0.834 | 0.825 | 0.782 | 0.751 | 0.77 | 0.811 | EAS < EUR < AMR < SAS = AFR = IND | Increased risk of COVID-19 related mortality | 71 |

**Table 5:** Genetic variants related to comorbidities that are risk factors of COVID-19 susceptibility/severity. ^p^ Effect allele frequency in Indian population. ^q^ EAF (India) compared with EAF in other populations (IND – India, EUR – Europe, AMR – America, SAS – South Asia, AFR – Africa, EAS – East Asia. ‘=’ denotes no significant difference between EAF of Indian and the other population. (EA – effect allele, OA – other allele, Ref – references).

| rsid | Gene | EA | OA | IND <sup>p</sup><br>(EAF) | AFR<br>(EAF) | AMR<br>(EAF) | EAS<br>(EAF) | EUR<br>(EAF) | SAS<br>(EAF) | EAF comparison <sup>q</sup> | Effect | Ref |
| --- | --- | --- | --- | --- | --- | --- | --- | --- | --- | --- | --- | --- |
| rs2285666 | ACE2 | A | G | 0.459 | 0.207 | 0.326 | 0.536 | 0.239 | 0.487 | AFR < EUR < AMR < IND = SAS < EAS | Increased risk of developing hypertension, which might increase the risk for COVID-19 severity | 145 |
| rs4240157 | ACE2 | C | T | 0.31 | 0.541 | 0.288 | 0.035 | 0.343 | 0.276 | EAS < SAS = AMR = IND < EUR < AFR | Increased risk of developing hypertension and Type 2 diabetes, which might increase the risk for COVID-19 severity | 50, 49 |
| rs4646188 | ACE2 | A | G | 0.938 | 0.997 | 0.968 | 1 | 0.864 | 0.939 | EUR < IND = SAS = AMR < AFR < EAS | Increased risk of developing hypertension, which might alter COVID-19 severity Increased risk of developing atrial fibrillation and cardioembolic stroke in diabetic patients | 50, 146 |
| rs2158083 | ACE2 | C | T | 0.24 | 0.172 | 0.245 | 0.005 | 0.336 | 0.188 | EAS < AFR < SAS < IND = AMR < EUR | Associated with changes in blood pressure, potentially affecting COVID-19 susceptibility and severity | 50 |
| rs1978124 | ACE2 | T | C | 0.276 | 0.098 | 0.29 | 0.005 | 0.456 | 0.208 | EAS < AFR < SAS < IND = AMR < EUR | Increased risk of developing hypertension, which might increase the risk for COVID-19 severity | 50, 130 |
| rs2074192 | ACE2 | T | C | 0.222 | 0.351 | 0.405 | 0.43 | 0.432 | 0.227 | IND < SAS < AFR < AMR < EAS < EUR | Increased risk of developing hypertension, potentially affecting COVID-19 susceptibility and severity | 49 |
| rs4646155 | ACE2 | T | C | 0.069 | 0.136 | 0.019 | 0.032 | 0.002 | 0.085 | EUR < AMR < EAS < IND = SAS < AFR | Increased risk of developing essential hypertension, thereby modulating COVID-19 susceptibility and severity, | 147 |
| rs2106809 | ACE2 | T | C | 0.545 | 0.913 | 0.687 | 0.477 | 0.751 | 0.509 | EAS < SAS = IND < AMR < EUR < AFR | Increased risk of developing essential hypertension, thereby modulating COVID-19 susceptibility and severity, | 148 |
| rs55790676 | ADAM17 | T | G | 0.075 | 0.081 | 0.134 | 0.02 | 0.184 | 0.096 | EAS < IND = AFR = SAS < AMR < EUR | Decreased total cholesterol levels and increased HDL levels, potentially affecting COVID-19 severity | 50 |
| rs2107538 | CCL5 | T | C | 0.281 | 0.451 | 0.225 | 0.322 | 0.161 | 0.309 | EUR < AMR < IND = SAS = EAS < AFR | Associated with type 2 diabetes and tuberculosis, which might affect COVID-19 susceptibility and severity | 149, 150 |
| rs1800796 | IL6 | C | G | 0.348 | 0.103 | 0.295 | 0.791 | 0.048 | 0.395 | EUR < AFR < AMR = IND = SAS < EAS | Increased risk of coronary artery disease, which might affect COVID-19 severity | 151 |
| rs3745264 | RAVER1 | A | C | 0.201 | 0.057 | 0.297 | 0.178 | 0.157 | 0.213 | AFR < EUR < EAS = IND = SAS < AMR | Increased risk of developing familial hypercholesterolemia | 152, 42 |

Comparing the risk allele frequencies of a variant between multiple populations can help gain insight into relative genetic predisposition to the associated condition at the population level [42, 43]. In the absence of case-control studies, determining the number of risk variants for different populations might help understand the cumulative genetic effect, hence simplifying the comparison. Here, we applied this approach to ‘COVID-19 susceptibility’, ‘COVID-19 severity’ and ‘COVID-19 related mortality’ categories. Risk conferring variants were assigned to the populations where their frequencies were either highest or lowest. For each population, the number of variants with the highest risk allele frequency and lowest risk allele frequency was counted (Supplementary File 1, Supplementary Table 3). In the ‘COVID-19 susceptibility’ category, the number of variants with the highest risk allele frequency was less than the variants with the lowest risk allele frequency in the IND population. A similar trend was observed in EAS and AFR populations. However, an opposite trend was observed in EUR and AMR populations (Fig. 1A). In IND and AFR populations, COVID-19 severity-related variants with the highest risk allele frequencies were more than the lowest frequency variants, while in EAS, EUR, and AMR populations this was found to be opposite (Fig. 1B). The number of variants associated with COVID-19 related mortality and having the highest risk allele frequency was less than the variants with the lowest risk allele frequency in IND, EAS, and AFR populations, while in SAS and AMR only the highest frequency variants were observed (Fig. 1C).

**Figure 1.**
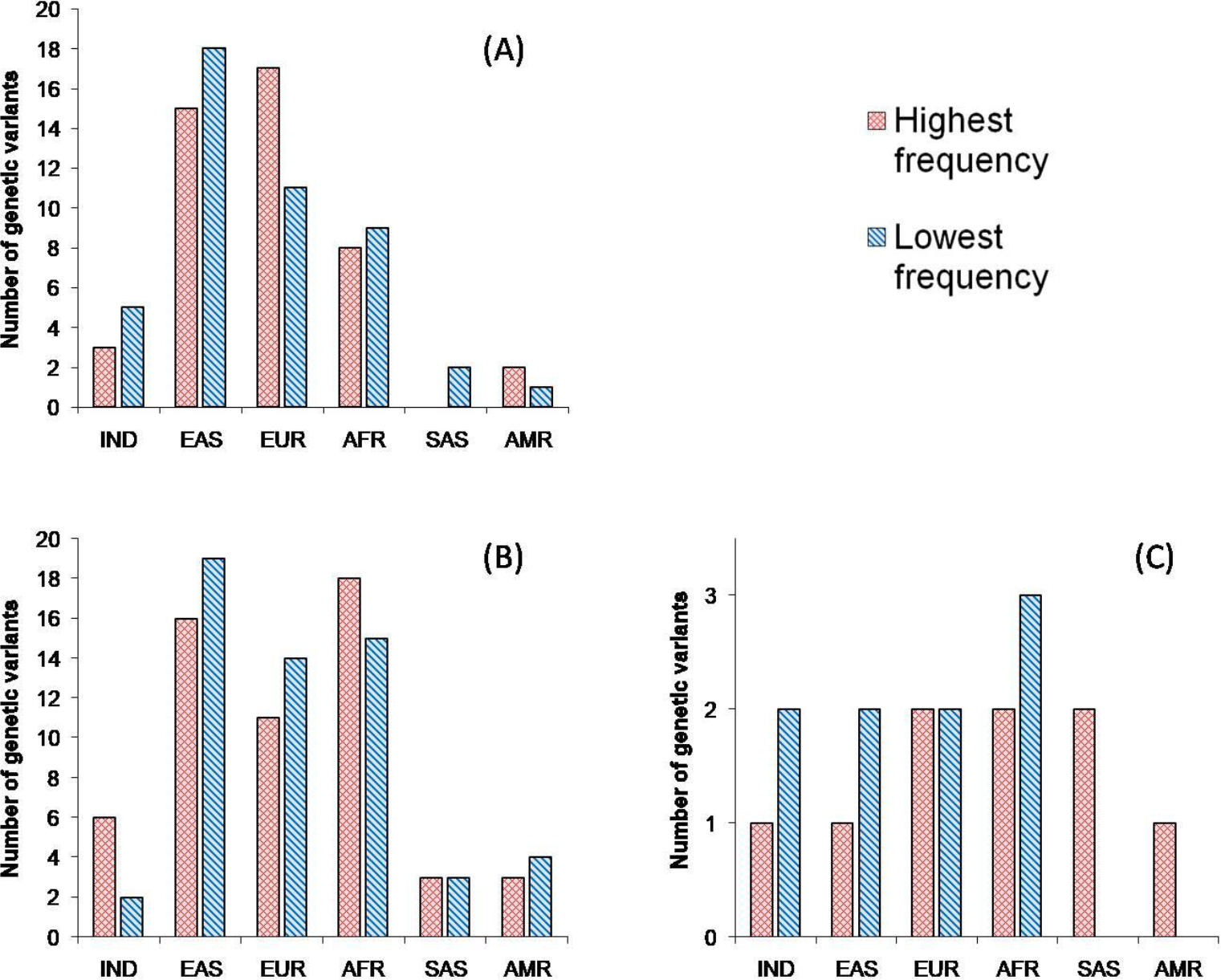
Comparison of populations based on the number of variants with highest / lowest risk allele frequency related to (A) Susceptibility to COVID-19, (B) COVID-19 severity, and (C) COVID-19 related mortality or fatal outcomes. IND – India, EUR – Europe, AMR

## 4 DISCUSSION

### 4.1 COVID-19 susceptibility

SARS-CoV-2 gains entry into human cells by binding its spike protein to ACE2 receptors. ACE2, a part of the renin-angiotensin-aldosterone system (RAAS), is crucial in regulating blood pressure and maintaining electrolyte balance in the body, physiology of the heart, kidneys, and lungs [44, 45]. Genetic variations in ACE2 are known to be associated with its increased expression levels in COVID-19 patients. These variants affect ACE2 transcription/translation and also alter binding affinity to SARS-CoV2 spike protein, thereby affecting the susceptibility to SARS-CoV-2 infection [46, 47]. Six ACE2 polymorphisms (rs2285666 [A], rs4240157 [T], rs4646188 [G], rs2158083 [T], rs1978124 [C] and rs233574 [T]), which are known to be associated with increased ACE2 expression, were found to have allele frequencies significantly different in Indian population as compared to others [48, 49, 41, 50, 51, 52]. Their effect allele frequencies in Indian population are 0.459, 0.69, 0.062, 0.76, 0.724 and 0.176, respectively (Table 1). For four of these variants, effect allele frequencies in EAS are highest suggesting that the EAS population might be predisposed to an increased susceptibility to COVID-19. In addition, the frequencies of rs1996225 [T] and rs4060 [A] of CA5BP1 gene and rs1893217 [G] of PTPN2 gene associated with increased expression of ACE2 are also highest in EAS. IND population has intermediate frequencies for most of these variants indicating moderate susceptibility, while EUR has the lowest frequencies indicating reduced susceptibility.

ACE (angiotensin-converting enzyme) is a vital part of the vascular system. It negatively regulates ACE2 and causes vasoconstriction. Alu Insertion in the ACE coding region inhibits ACE expression, while deletion promotes ACE expression. Increased ACE2 expression is observed when homozygous Alu insertion is present in the ACE gene and vice-versa. rs4343 is in high LD with this Alu element and thus used as its tag SNP [53]. The A and G alleles of rs4343 correspond to Alu insertion and deletion, respectively [54]. Thus A allele signifies reduced ACE expression and consequently increased ACE2 expression, potentially leading to increased SARS-CoV-2 susceptibility. In Indians, the frequency of the A allele (0.604) is significantly higher than Europeans (0.435) but lesser than other populations (Table 1).

On infection with SARS-CoV-2, the ADAM17 shedding is activated, leading to the release of ACE2 from the cell membrane into serum – thus inhibiting the virus infection. But, this effect would be compromised, if the infection rate is high [55, 56, 57, 58]. Thus ADAM17 mediated ACE2 levels may modulate COVID-19 susceptibility. ADAM17 releases the precursor TNF-α from the membrane to its active form – which is a crucial player in triggering an inflammatory response, leading to cytokine storm and is associated with severe lung damage in COVID-19 patients [59,60]. Thus, ADAM17 might be playing role in ACE2 mediated COVID-19 susceptibility and TNF-α mediated COVID-19 severity. rs55790676 [G] and rs1524668 [C] associated with decreased ADAM17 expression have high frequencies in IND population (0.925 and 0.403, respectively), lesser only than EAS, indicating increased susceptibility of COVID-19 in IND population.

FURIN is an endoprotease that is implicated in cleaving the SARS-CoV2 spike protein at the S1/S2 site and this cleavage is crucial for viral entry into pulmonary cells [61]. Increased FURIN expression affects monocyte adhesion and transendothelial migration [62] that can lead to tissue damage which is seen in severe COVID-19 patients [63]. In this study, three genetic variants that affect FURIN expression were identified whose allele frequencies are significantly different in the IND population. rs4702 [A] and rs4932178 [T], present in FURIN coding region, are associated with increased FURIN expression [64, 41]. It was also found that rs4702 [A] is associated with increased SARS-CoV2 infection [65]. The frequency of rs4702 [A] in IND (0.499) is one of the lowest suggesting relatively lesser COVID-19 susceptibility in the IND population. The frequency of rs4932178 [T] in the IND population (0.276) is intermediate as compared to others, indicating moderate COVID-19 susceptibility in IND. rs17514846 [A] is associated with the increased FURIN expression in vascular endothelial cells, where it affects monocyte adhesion and transendothelial migration [62]. Recruitment of monocytes and their transendothelial migration can lead to tissue damage which is seen in severe COVID-19 patients [63, 66]. The frequency of rs17514846 [A] in IND (0.371) is significantly higher than EAS and AMR populations and lesser than EUR (Table 3).

IFITM-family proteins are major players in antiviral responses [67]. Severe inflammation and enhanced activation of natural killer cells are observed in influenza-infected IFITM3 deficient mice [68]. IFITM3 is up-regulated in SARS-CoV2 infected lung epithelial cells of COVID-19 patients [69], and IFITM3 seems to restrict SARS-CoV2 infection by modulating the cell membrane properties [70]. This indicates that IFITM3 might be modulating susceptibility to SARS-CoV2 infection. rs11246066 [A] allele is associated with increased IFITM3 expression in whole blood and it is also an eQTL of other IFITM family genes in multiple tissues [71]. rs11246066 [A] allele frequency in the Indian population (0.189) is significantly greater only than the African population. rs9666637 [T] and rs7931303 [G] are associated with increased IFITM2 expression. rs9666637 [T] allele frequency in the Indian population (0.912) is highest among all populations, while rs7931303 [G] allele frequency in the Indian population (0.398) is higher only than AFR. Considering the significance of the IFITM family, Indian population might be less susceptible to COVID-19 due to rs9666637 [T].

CTSL gene is involved in many biological processes such as antigen processing and presentation, proteolysis, macrophage apoptotic process, and immune response. It is associated with many diseases such as cancer, diabetes, kidney failure, and viral infections [72]. It regulates viral entry into host cells and is up-regulated during COVID-19 infection suggesting its potential role in COVID-19 susceptibility and severity [72, 73]. rs3128509 [G] is associated with decreased CTSL expression in various tissues [41] and its frequency is significantly lesser in Indians (0.809) compared to African and East Asian populations but greater than European and American populations. This suggests that the Indian population might be more susceptible to COVID-19 than European and American populations but lesser than African and East populations.

Progesterone acts as a potent immunomodulator in regulating innate and adaptive immune responses and vascular system functions. Progesterone receptors (PGRs) are intracellular receptors that on binding to progesterone or its derivative regulate target gene expression. An Alu insertion in intron 7 of the PGR gene reduces transcript stability, consequently affecting PGR protein structure and function [74]. As a result of this insertion, PGR modulated differential expression of ACE2 may potentially affect SARS-CoV2 infection susceptibility. This Alu insertion is in complete LD with rs1042838. A and G alleles of rs1042838 correspond to Alu insertion and deletion, respectively [53]. The frequency of rs1042838 [A] in IND (0.078) is significantly lesser than AMR (0.137) and EUR (0.179) populations while greater than AFR (0.006) and EAS (0.01) populations.

TMPRSS2 is a serine protease that plays an important role in SARS-COV2 entry into the host cell. The viral spike (S) protein on binding to ACE2 on the host cell is cleaved at its S1/S2 position by TMPRSS2 thus facilitating viral entry into the host cell [75]. Increased SARS-CoV2 infection correlates with increased TMPRSS2 expression [76]. Eleven polymorphisms (rs35477708 [GA], rs468259 [C], rs363981 [T], rs1638369 [T], rs469288 [G], rs468699 [C], rs462698 [A], rs8134378 [G], rs467375 [A], rs4818239 [C], rs35899679 [A]) whose allele frequencies are significantly in Indian population are associated with increased TMPRSS2 expression [77, 64, 47, 41]. Except for rs8134378 [G], frequencies of remaining variants are highest in EUR and lowest in EAS, suggesting EUR might be at increased susceptibility of COVID-19 while EAS at lowest. Their frequencies in IND are intermediate thus potentially conferring moderate susceptibility in the IND population.

Zinc deficiency is associated with increased COVID-19 susceptibility in the Asian population [78]. Among the genetic variants associated with decreased zinc levels (rs2120019 [C], rs1532423 [G], rs4826508 [C]) and thereby increased susceptibility to COVID-19, rs1532423 [G] from CA1 gene and rs4826508 [C] from the NBDY gene have the lowest frequencies in IND (0.494 and 0.273, respectively), which might be conferring reduced susceptibility to COVID-19 in Indian population. However, rs2120019 [C] from the PPCDC gene might be having an opposite effect as its frequency in IND (0.62) is highest.

### 4.2 Immune response to SARS-CoV2 infection

IFIH1 gene encodes a protein that recognizes the viral RNA and initiates immune and proinflammatory responses. rs1990760 [T], which is present in the IFIH1 coding region, is associated with autoimmune disorders and protects against viral infection [79]. T allele frequency in Indians is 0.516, second only to Europeans (Table 2). Thus, this might be conferring increased protection to the Indian population against viral infections, including SARS-CoV2.

The chemokines CCL5, CCL4, and CCL3 play an important role in inflammation where they recruit immune cells [80, 81]. They are expressed during respiratory viral infections, and their expression levels are associated with disease severity [82, 83, 84]. In severely affected COVID-19 patients, their expression levels are significantly high, leading to liver toxicity and kidney failure, which are the most common complications associated with COVID-19 infection [82, 83, 84, 85]. rs2526327 [A] in RDM1 gene, rs1994182 [G] in MMP28 gene, rs3826404 [G] in MMP28 gene, rs2107538 [T] and rs3817655 [T] in CCL5 genes are associated with decreased CCL5 expression in whole blood [71] while rs4239252 [A] in TAF15 gene is associated with increased expression of CCL5 in whole blood [71]. All of these alleles are minor alleles in the Indian population. rs3817655, which lies in the CCL5 coding gene, is also associated with SARS-COV susceptibility [42]. The frequency of rs3817655 [T] allele in the Indian population (0.127) is least as compared to other populations.

### 4.3 COVID-19 severity

IGFBP2 is involved in the regulation of insulin-like growth factor receptor signaling pathways, and its expression level is used as a marker for many cancers. The intronic variant rs2270360 [A] in the IGFBP2 gene was found to be associated with increased COVID-19 severity [86]. The risk allele [A] frequency in the Indian population is 0.927 which is significantly higher than other populations, indicating the predisposition of the Indian population to the severity of the disease.

Low calcium levels in the serum are associated with COVID-19 severity, and maintaining proper calcium is a prerequisite to managing COVID-19 in the initial infection stage. RYR3 is a part of the calcium channel complex and releases calcium for intracellular activities [87]. rs2229117 [G] in the RYR3 gene is shown to be associated with increased COVID-19 severity [86]. G allele frequency in the Indian population (0.913) is higher than American and European populations but lesser than African and East Asian populations (Table 3). Thus this variant might be conferring a moderate risk of COVID-19 severity in the Indian population.

CCL2 and CCR1 play major roles in immune and inflammatory responses. CCL2 is a chemo-attractant cytokine that modulates the recruitment of monocytes, T cells, B cells, natural killer cells, macrophages, and dendritic cells [88]. Increased CCL2 levels are observed in severe COVID-19 patients as compared to less severe COVID-19 patients [89]. CCR1 is a chemokine receptor that is strongly expressed in monocytes and macrophages and promotes their infiltration into the lungs in severe COVID-19 cases [90]. rs1024611 [G] and rs1015164 [G] alleles are associated with increased CCL2 expression [91] and increased CCR1 expression in whole blood [63], respectively. rs1024611 [G] frequency in Indians (0.316) is significantly greater than only the African population, while rs1015164 [G] frequency in Indians (0.753) is significantly greater than only the European population (Table 3).

MX1 is known to be implicated in the generation of protective immune responses against influenza infection [92]. Increased MX1 expression in the nasopharynx of COVID-19 patients is observed to be proportional to SARS-CoV2 viral load. However, its expression seems to reduce with increased age, suggesting inadequate antiviral immune response in older adults causing severe COVID-19 [92]. These results suggest that increased expression of MX1 might be restricting the COVID-19 severity. Six polymorphisms (rs35477708 [G], rs468259 [G], rs363981 [G], rs1638369 [G], rs469288 [A], rs468699 [T], rs462698 [G]) associated with decreased MX1 expression [41, 47] might be increasing risk of COVID-19 severity. Effect allele frequencies of each of these variants are highest in EAS and lowest in the EUR population. This indicates that the EAS population might be vulnerable to increased risk of COVID-19 severity, while EUR and IND populations might be at reduced and moderate risk, respectively.

TNF encodes a proinflammatory cytokine secreted by macrophages is a major player contributing to the development of cytokine storms. rs1800630 [A] in the TNF gene is associated with an increased risk of developing acute respiratory distress syndrome (ARDS) [93]. The frequency of risk allele [A] in the Indian population (0.283) is highest as compared to other populations, suggesting that the allele might be conferring increased risk of COVID-19 severity to the Indian population.

AHSG levels are critical for regulating macrophage deactivation. Decreased AHSG leads to impaired macrophage deactivation thus causing excess pro-inflammatory cytokine release in SARS-COV infection. rs2248690 [T] is associated with increased AHSG levels and decreased SARS-COV infection severity [94]. T allele frequency in the Indian population is 0.17 which is the least as compared to other populations, indicating that the Indian population might be at an increased risk of COVID-19 severity.

rs10735079 in OAS gene cluster (OAS1, OAS2, OAS3) is associated with antiviral mechanisms by producing 2′,5′-oligoadenylate (2-5A), which is a mediator in the antiviral process. This mediator activates RNaseL which degrades double-stranded RNA of corona virus [95]. rs10735079 [A] allele is associated with an increased risk of severe COVID-19. rs10735079 [A] allele corresponds to lower OAS1 activity and higher odds of severe COVID-19 [95]. A allele frequency in the Indian population (0.698) is lower than African, American, and East Asian populations but greater than Europeans, suggesting that the Indian population might be at a reduced risk of COVID-19 severity.

Decreased levels of E-selectin and IL-3Ra proteins and increased levels of B3GN2 and C1GLC proteins are associated with increased COVID-19 severity [96]. rs2519093 [T] is associated with decreased E-selectin and IL-3Ra protein levels and increased B3GN2 and C1GLC protein levels. Thus, rs2519093 [T] might be increasing COVID-19 severity through modulating these protein levels. T allele frequency in the Indian population (0.128) (Table 3), is greater than only the African population, suggesting that Indian population might be at a reduced risk of COVID-19 severity.

Vitamin D deficiency is associated with COVID-19 positivity and severity [97, 98]. GC gene encodes a protein that binds to vitamin D and transports it to targeted tissues. rs1155563 [C] allele, that is present in the GC gene, is associated with decreased serum vitamin-D levels [99], and its frequency in the Indian population (0.29) is greater than that of African, American, and European populations (Table 3). This suggests that rs1155563 [C] might be increasing COVID-19 susceptibility and severity in the Indian population through reduced vitamin-D levels.

### 4.4 COVID-19 related mortality or fatal outcomes

Kim YC and Jeong BH [71] found the frequencies of T allele of rs2074192 in the ACE2 gene, and G allele of rs2298659 in the TMPRSS2 gene to be positively correlated with case fatality rate. Indians have lowest frequency of rs2074192 [T] (0.222) compared to Africans (0.351), Americans (0.405), East Asians (0.43) and Europeans (0.432). But they have highest frequency of rs2298659 [G] (0.834) compared to Africans (0.825), Americans (0.782), Europeans (0.77) and East Asians (0.751). Indians have lesser frequency of G allele of rs6598045 in the IFITM3 gene (0.159), which is associated with decreased case fatality rate, compared to Africans (0.299) and Americans (0.222).

DES/SPEG genes code for muscle-specific proteins and mutations in these genes are involved with cardiomyopathies [100, 101]. Acute damage to the heart is a common complication in COVID-19 patients. A allele of rs71040457 that is located downstream of the DES and upstream of the SPEG genes was found to be associated with an increased risk of COVID-19 mortality in the white British ancestry population [102]. While East Asians have the lowest frequency of this allele (0.177), Indians have a lesser frequency (0.276) compared to European (0.376), American (0.385), and African (0.939) populations.

SLC39A10 gene plays an important role in mediating immune cell homeostasis. It has been reported to facilitate anti-apoptotic signaling during early B-cell development, modulate B-cell receptor signal strength, and control macrophage survival. rs113892140 located in this gene is associated with an increased risk of COVID-19 mortality [102]. Risk allele [A] frequency in the Indian population is 0.078 (Table 4), which is significantly greater than only the European population (0.046). Its frequency is highest in the African population (0.256).

### 4.5 COVID-19 related comorbidities

Twelve polymorphisms associated with comorbidities of COVID-19 were also analyzed (Table 5). Hypertension, coronary artery disease, type 2 diabetes are among the major comorbidities of COVID-19 severity. rs2074192 [T] is associated with an increased risk of developing hypertension [49]. T allele frequency in the Indian population (0.222) is least as compared to other populations.

## 5 CONCLUSION

The potential role of several polymorphisms in COVID-19 susceptibility and severity suggests that host genetics plays an important role in the pathology and progression of the infection. We found multiple genetic variants that might be affecting the COVID-19 susceptibility, immune response, severity, and mortality. Some of these variants were observed in genetic association studies, while others were found to be relevant based on gene regulation and signaling pathways.

COVID-19 positivity rate in India is one of the least in the world despite India being the second most populated nation [154, 155]. Due to the high population density in India, the spread of an airborne pathogen can be limited only to a certain extent [103]. Hence, a low COVID-19 positivity rate in India might be due to the lower genetic predisposition of Indians to COVID-19 susceptibility. The fatality rate of COVID-19 in India was relatively high during the peak phases of the first and second waves. However, when the peaks subsided the fatality rate reduced significantly. Inadequate medical facilities seem to be one of the major reasons which caused preventable deaths during these periods. [104]. Thus, the role of genetics in COVID-19 severity risk in the Indian population remains unclear. More detailed studies are warranted to confirm the COVID-19 related relevance of the variants discussed in this article. The current study will act as a good source to shortlist variants/genes for conducting genetic association studies to assess COVID-19 susceptibility, its severity, and mortality.

## Data Availability

All data produced in the present study are available upon reasonable request to the authors

